# An exposome-wide investigation of 2923 Olink proteins with non-genetic factors in Chinese adults

**DOI:** 10.1101/2024.10.23.24315975

**Authors:** Andri Iona, Baihan Wang, Jonathan Clarke, KaHung Chan, Maria G. Kakkoura, Charlotte Clarke, Neil Wright, Pang Yao, Mohsen Mazidi, Pek Kei Im, Maryam Rahmati, Christiana Kartsonaki, Sam Morris, Hannah Fry, Iona Y Millwood, Robin G Walters, Yiping Chen, Huaidong Du, Ling Yang, Daniel Avery, Dan Valle Schmidt, Feifei Li, Canqing Yu, Dianjianyi Sun, Jun Lv, Michael Hill, Liming Li, Robert Clarke, Derrick A Bennett, Zhengming Chen, China Kadoorie Biobank Collaborative Group

## Abstract

**Background:** Previous studies in European populations have identified a large number of genetic variants affecting plasma levels of Olink proteins, but little is known about the non-genetic factors influencing plasma levels of proteins, particularly in Chinese populations.

**Methods:** We measured plasma levels of 2,923 proteins, using Olink Explore platform, in 2,006 participants in the China Kadoorie Biobank. Linear regression analyses were used to assess the cross-sectional associations of individual proteins with 37 exposures across multiple domains (e.g. socio-demographic, lifestyle, environmental, sample processing, reproductive factors, clinical measurements, and health-related indices), adjusted for potential confounders and multiple testing. These were further replicated and compared with similar analyses in Europeans.

**Results:** Overall 31 exposures were associated with at least one protein, with age (n=1,154), sex (n=827), BMI (n=869) showing the highest number of associations, followed by frailty index (n=597), SBP (n=479), RPG (n=387), ambient temperature (n=292), and HBsAg-positivity (n=282), with diet and physical activity showing little associations. Likewise, of the 2,923 proteins examined, 65% were associated with at least one exposure, with three proteins (CDHR2, CKB, and PLAT) showing the largest number of associations with baseline characteristics (n=14). The patterns of associations differed by sex, chiefly due to differences in lifestyle and reproductive factors. Over 90% of proteomic associations with key exposures in the current study were replicated in the UK Biobank.

**Conclusions:** In Chinese adults, the exposome-wide assessment of Olink proteins identified a large number of associations with a wide range of exposures, which could inform future research priorities and analytic strategies.

## Introduction

Deciphering the human proteome could enhance our understanding of human health and disease.^1^ Plasma levels of proteins, which are secreted or leaked from cells or organs, may be affected by various genetic and non-genetic factors relevant to human health.^2^ Investigating the relationships between plasma proteins and different exposures could improve our understanding of human biology and inform research strategies.

Traditionally, mass spectrometry has been used to measure plasma protein levels,^3–5^ but studies using this method are typically constrained by their small sample sizes and low breadth of coverage.^2^ In contrast, advances in affinity-based technologies (e.g. Olink and SomaScan) have made it possible to leverage proteomics in large-scale population and clinical studies, thus allowing for a more comprehensive investigation of plasma proteins and their relationships with a variety of factors and health conditions.^2,6,7^ In particular, the Olink platform which utilises antibodies as reagents to bind target proteins, has been widely used in epidemiological research due to its high sample throughput and assay specificity.^6^ Recently, the Olink Explore 3072 platform was used to measure 2,923 plasma proteins in 54,219 participants in the UK Biobank,^8^ leading to many novel findings linking protein abundance with a range of demographic and clinical exposures and genetic factors.^8^ It also replicated some well-established associations, such as the associations between sex and LEP, and between age and GDF15.^6–11^ Another affinity-based platform, SomaScan,^7^ has also been used by other studies,^12,13^ but analysis of such studies focused mainly on genetic analyses.

Most previous proteomics studies investigated only a few pre-selected exposures, without simultaneously considering a broader range of factors that might be associated with the plasma proteome. Moreover, few studies have investigated associations of plasma proteins with composite indices (e.g. frailty) derived from various measures that may reflect general lifestyle and health,^14–17^ which could be useful for population-level screening and disease prevention. Furthermore, previous studies typically focused on the discovery of protein quantitative trait loci (pQTLs).^8,12,13^ While useful for downstream analyses such as Mendelian Randomisation to inform drug development,^2^ they do not contribute important insights into the roles of non-genetic factors in influencing levels of circulating proteins. Finally, there is evidence that protein concentrations can vary across different populations,^18^ which could further affect their associations with exposures. However, the lack of diversity remains an issue in proteomics, as the majority of proteomic studies in large-scale population-based cohorts have been conducted in European populations. Our previous study in the China Kadoorie Biobank (CKB) identified 27 proteins associated with body mass index (BMI), but this was conducted in a small sample of 628 participants and with an older Olink panel covering 92 proteins.^19^

To fill the evidence gap, the present study aims to use an “exposome-wide” approach to (1) comprehensively explore the exposure profiles of ∼3,000 Olink proteins in 2,000 Chinese adults in the CKB; (2) assess the consistency of proteomic associations between the Chinese and European populations; and (3) identify priorities for future research. We also conducted parallel analyses in the accompanied report on ∼7,000 proteins measured by the SomaScan platform in the same sample.^20^

## Methods

### Study population and design

The CKB is a prospective cohort study of >512,000 adults who were recruited from 2004 to 2008 in 10 geographically diverse areas.^21,22^ At baseline, detailed information was collected from all participants using laptop-based questionnaires, including socio-demographic characteristics, medical history, and lifestyle habits, in addition to physical measurements including body composition and blood pressure. Non-fasting (with time since the last meal recorded) blood samples were also collected, processed, aliquoted, and then stored in liquid nitrogen for future unspecified research use. All participants provided written informed consent.

The present study was based on a case-subcohort study of IHD involving 1951 cases and 2026 subcohort participants who had no prior history of cardiovascular disease and no reported use of lipid-lowering medications.^23^ The subcohort participants were randomly selected from a population subset of 69,353 genotyped participants who were unrelated to each other.

### Proteomic assays

The plasma samples of all 3,977 participants collected at baseline were assayed by the Olink Explore 3072 platform that targets 2,923 unique proteins in four separate panels. The samples were retrieved from liquid nitrogen, thawed, and aliquoted into 96-well plates (including 8 wells per plate for external QC samples). They were then shipped separately to Olink laboratories in Uppsala, Sweden (first batch; 1,472 proteins) and Boston, USA (second batch; 1,469 proteins) for proteomic profiling. The final measurements of protein levels were provided in the arbitrary Normalized Protein eXpression (NPX) unit on a log2 scale. Six proteins were replicated in four panels and showed high correlations across panels (r>0.8), so only one measure was kept for each replicated protein. Details of individual proteins are shown in **eTable 1**. Further details on proteomic assays in CKB have been previously described.^23,24^

### Selected baseline characteristics

We selected 37 exposures across 6 broad categories **(eTable 2)**, covering demographics (e.g., age, sex, study area), lifestyle habits (e.g., alcohol, smoking, diet, physical activity), environmental factors (e.g., outdoor temperature, fasting time), health and wellbeing (e.g., prior disease and mental health), clinical measures (e.g., BMI, SBP, RPG) and female reproductive factors (e.g., age at menarche, age at menopause, parity). We also derived a healthy lifestyle index (ranging from 0 to 5, with a higher score indicating a healthier lifestyle) based on smoking, alcohol intake, physical activity, dietary habits, and body shape.^15–17^ Similarly, a frailty index based on an accumulation of age-related deficits was computed considering medical conditions (based on self-reports of diagnosis by a doctor or physical measurements), symptoms, signs, and physical measurements, of which the procedure is described in a previous publication.^14,25^

### Statistical analyses

The main analyses were conducted in 2006 subcohort participants only (after excluding 20 participants with missing data on outdoor temperature) and separately by sex.

The prevalence or mean values of selected baseline variables were standardised to the age (5-year groups), sex, and study area. Plasma protein levels were standardized (i.e. values divided by their SD) and analysed as continuous variables. Linear regression was used to examine the associations of different baseline characteristics with protein biomarkers, adjusting for age, age^2^, sex, study area, fasting time, fasting time^2^, outdoor temperature, outdoor temperature^2^, and plate ID.

As many proteins and exposures are correlated with each other, we followed the approach by Gadd et al. (2023) to correct for multiple testing.^26^ We performed principal component analysis for 2,923 unique proteins and found 834 components explaining 90% of the cumulative variance (**eFigure 1 and eTable 3**). Similarly, we performed principal component analysis for 32 exposures measured in both females and males and found 21 components explaining 90% of the cumulative variance. Taking into account 834 components for proteins and 26 components for exposures (21 + 5 reproductive factors measured only in females), we derived a Bonferroni-adjusted p-value threshold: 0.05/(834□×□26)□=□2.305□×□10^−6^. This adjustment was applied across all linear regression models.

**Figure 1.**
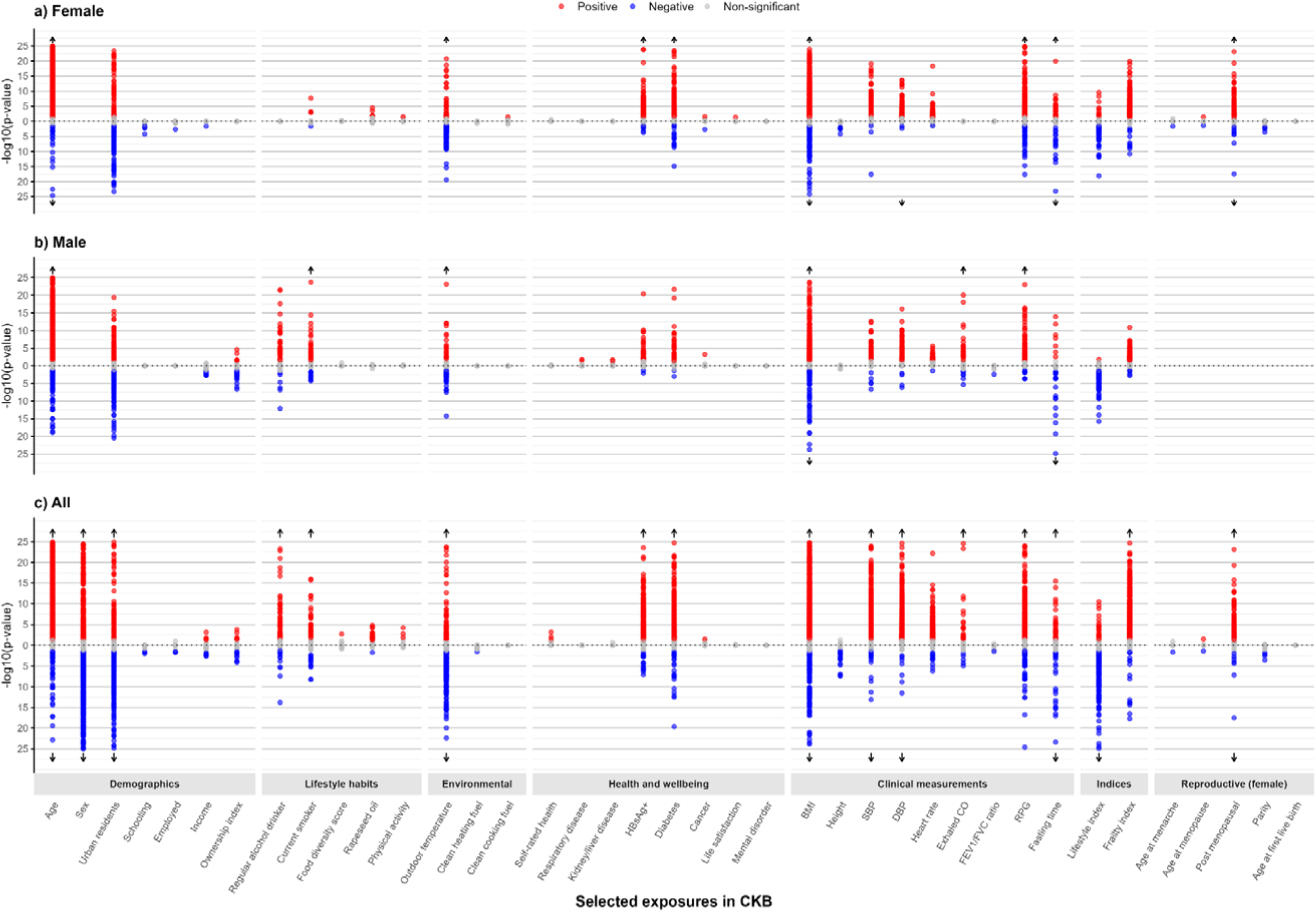
Exposure profiles with 2923 protein biomarkers in CKB, overall and by sex. Three Miami plots are presented: one for female-specific analysis, one for male-specific analysis, and one for overall analysis. The x-axis represents baseline characteristics grouped by category, while the y-axis shows the negative logarithm of the p-value (-log10 p-value) for the association between each exposure and protein biomarkers. Each dot represents the -log10 Bonferroni corrected p-value for these associations. For visualization purposes, -log10 p-values exceeding 25 are not displayed (indicated with arrow). Positive associations are shown in red, negative associations in blue, and non-significant associations in grey. Analyses are adjusted for age, age^2^, sex, study area, fasting time, fasting time^2^, outdoor temperature, outdoor temperature^2^ and plate ID, where appropriate. Abbreviations: BMI: Body mass index; CKB: China Kadoorie Biobank; CO: carbon-monoxide; DBP: Diastolic blood pressure; FEV1/FVC: Forced Expiratory Volume in 1 second / Forced Vital Capacity; HBsAg+: Hepatitis B virus surface antigen seropositive; RPG: random plasma glucose

We also undertook separate analyses of the same 2923 Olink proteins in approximately 35,000 white participants from the UK Biobank to replicate the main study findings in CKB, with the exclusion of participants with prior CVD or use of cholesterol-lowering medication.^8^

All statistical analyses were performed using R version 4.1.2^27^ and packages ‘tidyverse’, and ‘ggplot2’.

## Results

Among the 2,006 participants, the mean baseline age was 50.8 (SD 10.5) years, 62% of participants were female and the mean BMI was 23.9 (3.4) kg/m^2^ (**Table 1**). Overall, 15% of participants were regular alcohol drinkers (men: 37%; women 3%) and 25% (men: 63%; women: 2%) were current smokers. The prevalence of prior diseases was similar in males and females, with 8% of participants having respiratory disease, 2% having kidney/liver disease or tested sero-positive for HBsAg, and 6% having diabetes (self-reported or screen-detected).

**Table 1.**
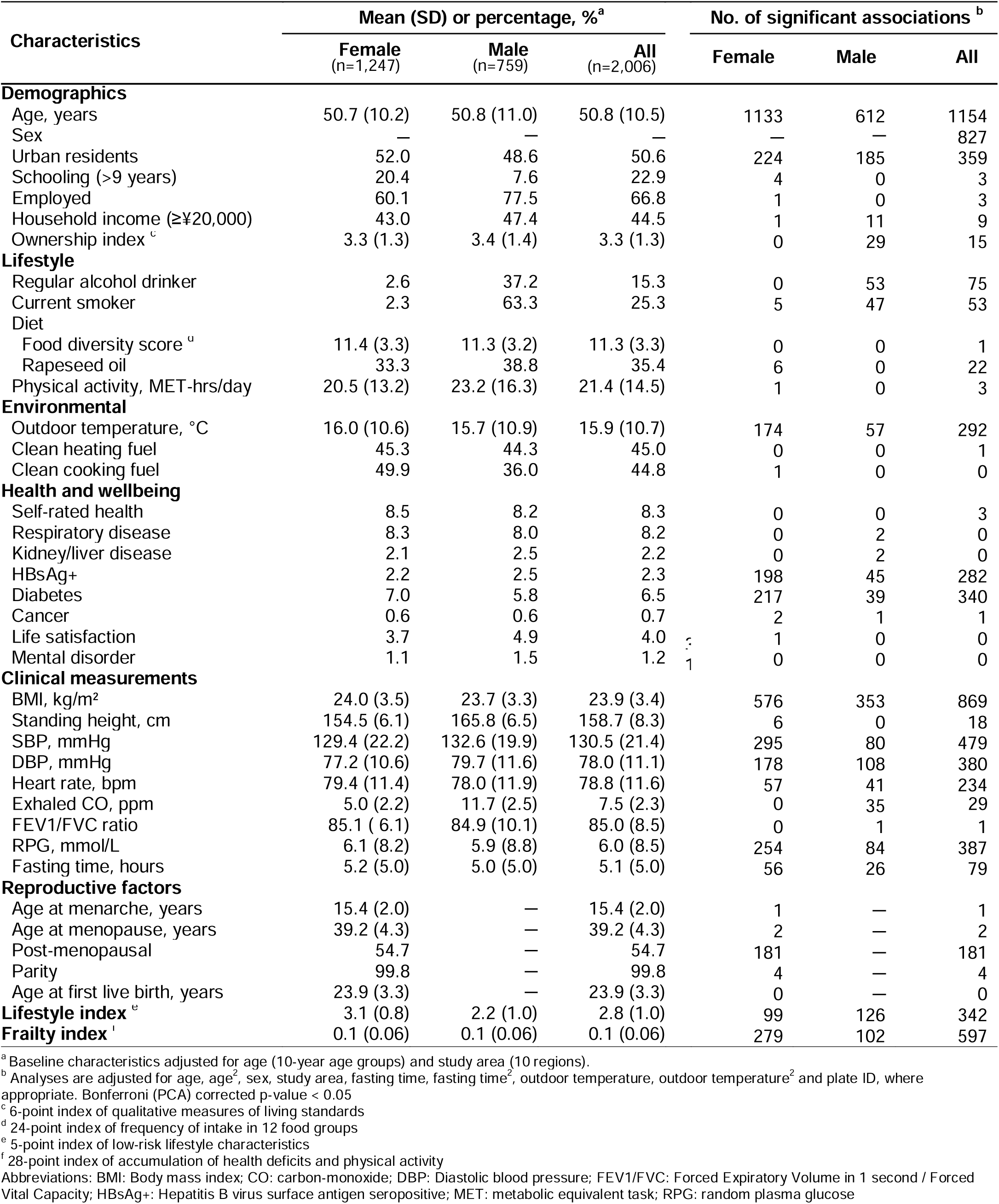
Baseline characteristics of participants and their associations with Olink protein biomarkers.

| Characteristics | Mean (SD) or percentage, %<sup>a</sup> |  |  | No. of significant associations<sup>b</sup> |  |  |
| --- | --- | --- | --- | --- | --- | --- |
|  | Female<br>(n=1,247) | Male<br>(n=759) | All<br>(n=2,006) | Female | Male | All |
| **Demographics** |  |  |  |  |  |  |
| Age, years | 50.7 (10.2) | 50.8 (11.0) | 50.8 (10.5) | 1133 | 612 | 1154 |
| Sex | — | — | — | — | — | 827 |
| Urban residents | 52.0 | 48.6 | 50.6 | 224 | 185 | 359 |
| Schooling (>9 years) | 20.4 | 7.6 | 22.9 | 4 | 0 | 3 |
| Employed | 60.1 | 77.5 | 66.8 | 1 | 0 | 3 |
| Household income (≥¥20,000) | 43.0 | 47.4 | 44.5 | 1 | 11 | 9 |
| Ownership index<sup>c</sup> | 3.3 (1.3) | 3.4 (1.4) | 3.3 (1.3) | 0 | 29 | 15 |
| **Lifestyle** |  |  |  |  |  |  |
| Regular alcohol drinker | 2.6 | 37.2 | 15.3 | 0 | 53 | 75 |
| Current smoker | 2.3 | 63.3 | 25.3 | 5 | 47 | 53 |
| Diet |  |  |  |  |  |  |
| Food diversity score<sup>d</sup> | 11.4 (3.3) | 11.3 (3.2) | 11.3 (3.3) | 0 | 0 | 1 |
| Rapeseed oil | 33.3 | 38.8 | 35.4 | 6 | 0 | 22 |
| Physical activity, MET-hrs/day | 20.5 (13.2) | 23.2 (16.3) | 21.4 (14.5) | 1 | 0 | 3 |
| **Environmental** |  |  |  |  |  |  |
| Outdoor temperature, °C | 16.0 (10.6) | 15.7 (10.9) | 15.9 (10.7) | 174 | 57 | 292 |
| Clean heating fuel | 45.3 | 44.3 | 45.0 | 0 | 0 | 1 |
| Clean cooking fuel | 49.9 | 36.0 | 44.8 | 1 | 0 | 0 |
| **Health and wellbeing** |  |  |  |  |  |  |
| Self-rated health | 8.5 | 8.2 | 8.3 | 0 | 0 | 3 |
| Respiratory disease | 8.3 | 8.0 | 8.2 | 0 | 2 | 0 |
| Kidney/liver disease | 2.1 | 2.5 | 2.2 | 0 | 2 | 0 |
| HBsAg+ | 2.2 | 2.5 | 2.3 | 198 | 45 | 282 |
| Diabetes | 7.0 | 5.8 | 6.5 | 217 | 39 | 340 |
| Cancer | 0.6 | 0.6 | 0.7 | 2 | 1 | 1 |
| Life satisfaction | 3.7 | 4.9 | 4.0 | 1 | 0 | 0 |
| Mental disorder | 1.1 | 1.5 | 1.2 | 0 | 0 | 0 |
| **Clinical measurements** |  |  |  |  |  |  |
| BMI, kg/m<sup>2</sup> | 24.0 (3.5) | 23.7 (3.3) | 23.9 (3.4) | 576 | 353 | 869 |
| Standing height, cm | 154.5 (6.1) | 165.8 (6.5) | 158.7 (8.3) | 6 | 0 | 18 |
| SBP, mmHg | 129.4 (22.2) | 132.6 (19.9) | 130.5 (21.4) | 295 | 80 | 479 |
| DBP, mmHg | 77.2 (10.6) | 79.7 (11.6) | 78.0 (11.1) | 178 | 108 | 380 |
| Heart rate, bpm | 79.4 (11.4) | 78.0 (11.9) | 78.8 (11.6) | 57 | 41 | 234 |
| Exhaled CO, ppm | 5.0 (2.2) | 11.7 (2.5) | 7.5 (2.3) | 0 | 35 | 29 |
| FEV1/FVC ratio | 85.1 (6.1) | 84.9 (10.1) | 85.0 (8.5) | 0 | 1 | 1 |
| RPG, mmol/L | 6.1 (8.2) | 5.9 (8.8) | 6.0 (8.5) | 254 | 84 | 387 |
| Fasting time, hours | 5.2 (5.0) | 5.0 (5.0) | 5.1 (5.0) | 56 | 26 | 79 |
| **Reproductive factors** |  |  |  |  |  |  |
| Age at menarche, years | 15.4 (2.0) | — | 15.4 (2.0) | 1 | — | 1 |
| Age at menopause, years | 39.2 (4.3) | — | 39.2 (4.3) | 2 | — | 2 |
| Post-menopausal | 54.7 | — | 54.7 | 181 | — | 181 |
| Parity | 99.8 | — | 99.8 | 4 | — | 4 |
| Age at first live birth, years | 23.9 (3.3) | — | 23.9 (3.3) | 0 | — | 0 |
| **Lifestyle index<sup>e</sup>** | 3.1 (0.8) | 2.2 (1.0) | 2.8 (1.0) | 99 | 126 | 342 |
| **Frailty index<sup>f</sup>** | 0.1 (0.06) | 0.1 (0.06) | 0.1 (0.06) | 279 | 102 | 597 |

<sup>a</sup> Baseline characteristics adjusted for age (10-year age groups) and study area (10 regions).<sup>b</sup> Analyses are adjusted for age, age<sup>2</sup>, sex, study area, fasting time, fasting time<sup>2</sup>, outdoor temperature, outdoor temperature<sup>2</sup> and plate ID, where appropriate. Bonferroni (PCA) corrected p-value < 0.05<sup>c</sup> 6-point index of qualitative measures of living standards<sup>d</sup> 24-point index of frequency of intake in 12 food groups<sup>e</sup> 5-point index of low-risk lifestyle characteristics<sup>f</sup> 28-point index of accumulation of health deficits and physical activity
Abbreviations: BMI: Body mass index; CO: carbon-monoxide; DBP: Diastolic blood pressure; FEV1/FVC: Forced Expiratory Volume in 1 second / Forced Vital Capacity; HBsAg+: Hepatitis B virus surface antigen seropositive; MET: metabolic equivalent task; RPG: random plasma glucose

Among 37 baseline characteristics examined, 31 were associated with at least one protein at the Bonferroni-adjusted threshold (**Table 1**, **Figure 1**). The three baseline characteristics that showed the largest number of associations with proteins were age (n=1154), sex (n=827) and BMI (n=869). Likewise, of the 2,923 proteins examined, 1900 (65%) were associated with at least one exposure, with three proteins (CDHR2, CK-BB, and PLAT) showing the largest number of associations with baseline characteristics (n=14), involving primarily demographic factors and clinical measurements (**Figure 2, eFigure 2**).

**Figure 2.**
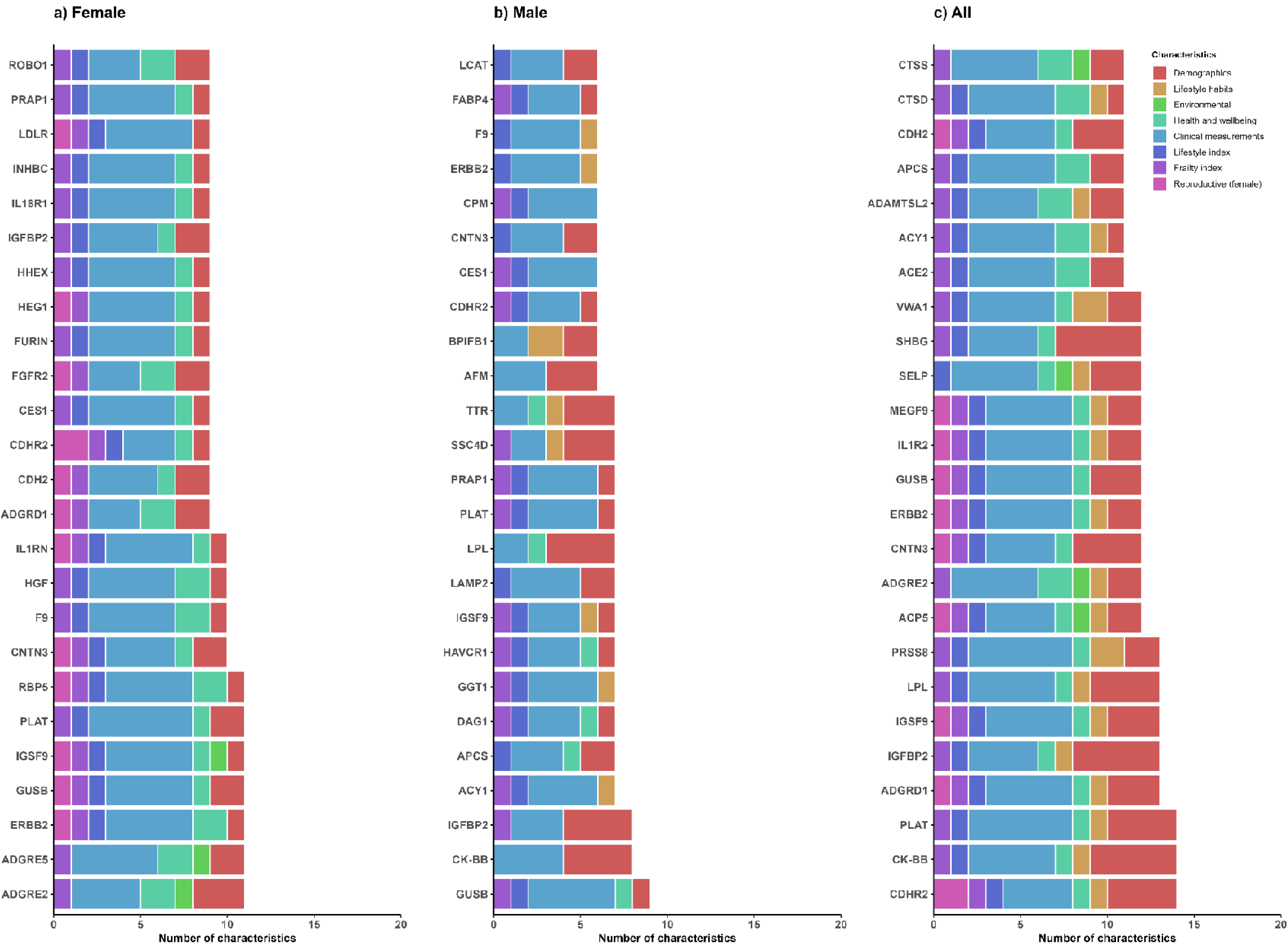
Exposure profiles by characteristics type of the top 25 protein biomarkers with most associations, overall and by sex. The bar plots show the number of baseline associated with the 25 most frequently associated protein biomarkers based on Bonferroni-corrected p-values. The analyses are presented separately for females, males, and overall. The x-axis represents the protein biomarkers, while the y-axis indicates the number of baseline characteristics associated with each protein. Bars are color-coded to represent different baseline characteristic groups. Analyses are adjusted for age, age^2^, sex, study area, fasting time, fasting time^2^, outdoor temperature, outdoor temperature^2^ and plate ID, where appropriate.

Of the 827 sex-related proteins (higher levels in females for 259 and in males for 568 proteins), the strongest associations were observed for LEP, XG and FSHB in females and for ACRV1, EDDMEB and INSL3 in males (**Figure 3 I.a**). Among the top 50 sex-related proteins, most were also associated with other exposures, chiefly age (e.g., FSHB, CGA, XG) and BMI (e.g., LEP, FABP4, CDHR2; **Figure 3 I.b**). Additionally, 77 sex-related proteins were not associated with any other exposures examined, of which 27 were uniquely associated with female sex (e.g., CSF3, MELTF, ITIH4) and 50 with male sex (e.g., EDDM3B, TEX101 and CRISP2; **eTable 4**).

**Figure 3.**
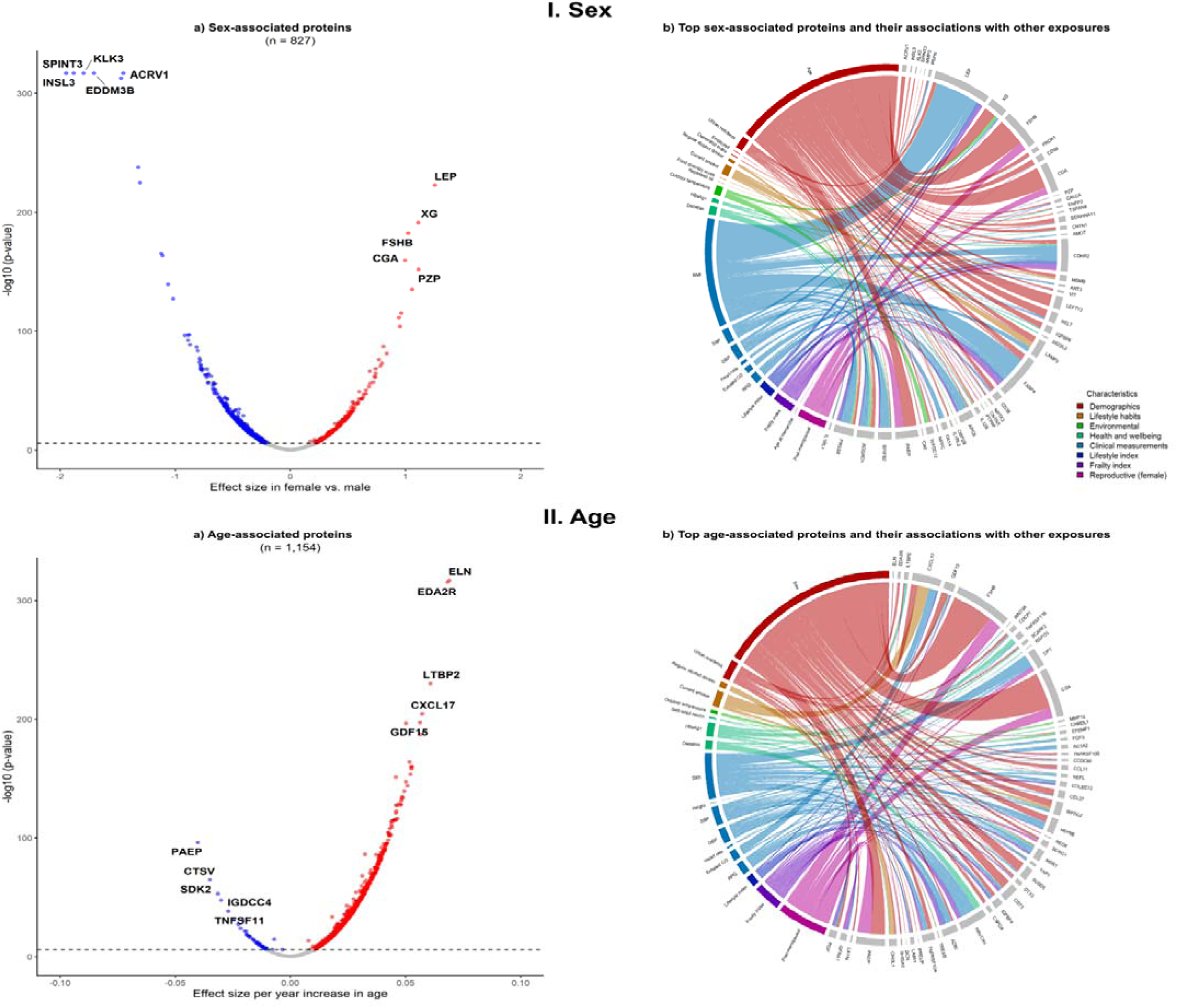
Sex- and age-associated protein biomarkers and their associations with other exposures. Figures a) and c) represent the associations of sex and age, respectively, with protein biomarkers. The x-axis represents the effect size of the association between sex or age and the protein biomarkers, while the y-axis indicates the –log10 p-value. Red dots denote positive Bonferroni corrected associations, blue dots denote negative Bonferroni corrected associations, and grey dots denote non-significant associations. Figures b) and d) illustrate the top sex-and age-associated protein biomarkers, respectively, and their associations with other exposures. The width of the ribbons is inversely proportional to the p-value, indicating the strength of the association (smaller p-values correspond to wider ribbons). The colors of the ribbons represent different baseline characteristic groups. The top protein biomarkers that are not associated with other exposures are not presented in the figure. Analyses are adjusted for age, age^2^, sex, study area, fasting time, fasting time^2^, outdoor temperature, outdoor temperature^2^ and plate ID, where appropriate. Abbreviations: BMI: Body mass index; CO: carbon-monoxide; DBP: Diastolic blood pressure; HBsAg+: Hepatitis B virus surface antigen seropositive; RPG: random plasma glucose

Of the 1,154 age-related proteins, the strongest positive associations were observed with EDA2R, ELN and LTBP2, and the strongest negative associations were with PAEP, CTSV and SDK2 (**Figure 3 II.a**). Among the top 50 age-related proteins, most were also associated with other exposures examined, chiefly sex (e.g., FSHB, CGA, PAEP) and BMI (e.g., DPT, HSPB6, FGF5; **Figure 3 II.b**). Additionally, 168 age-related proteins (e.g., ITGB5, IL17D, and TIMP4) were not associated with any other exposures (**eTable 4**). In sex-specific analyses, age was associated with 1,133 and 612 proteins in males and females, respectively (**Table 1; eFigure 3 I.a** & **II.b**). Among the 524 overlapping proteins in both sexes, nearly all (>95%) of the associations were directionally consistent, but the associations appeared stronger in females (r=0.62; **eFigure 4**). Among females, FSHB, ELN and EDA2R showed the strongest positive associations with age, while PAEP, SDK2 and CTSV showed the strongest negative associations (**eFigure 3 I.a**). Among males, EDA2R, ELN and LTBP2 showed the strongest positive associations, and EGFR, INSL3 and IGFBP3 showed the strongest negative associations (**eFigure 3 II.a**). Among females the top age-related proteins were predominantly associated with menopause, while among males they were mainly associated with clinical measurements (i.e., BMI, SBP, and RPG), exhaled CO and current smoking status in males (**eFigure 3 I.b** & **3 II.b**). Additionally, 394 and 352 age-related proteins in females and males, respectively, were not associated with any other exposures examined (**eTable 4**).

**Figure 4.**
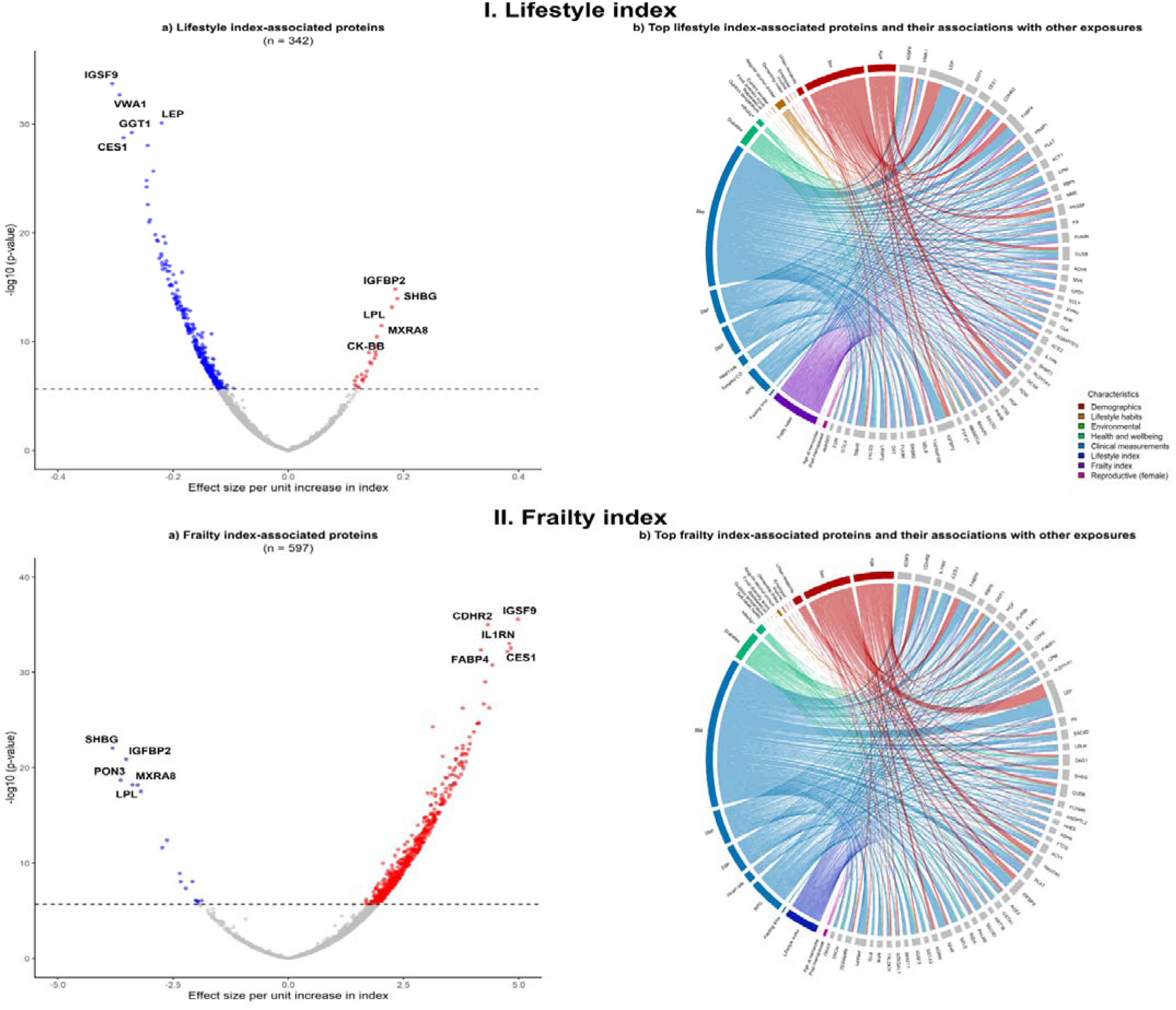
Lifestyle and frailty indices-associated protein biomarkers and their associations with other exposures. Symbols and conventions as in Figure 3.

In overall analyses, regular alcohol consumption and current smoking were associated with 75 and 53 proteins (4 overlapping), respectively, with MAMDC4, CHI3L1 and VWA1 most strongly associated with alcohol drinking and CXCL17, LAMP3 and ALPP most strongly associated with smoking (**Table 1, eFigure 5**). In separate analyses among men, most of these protein associations with alcohol (n=53) and smoking (n=47) were significant (**eFigure 6**). Overall, outdoor temperature was associated with 292 proteins (e.g., SNED1, SPINK6 and LRP1), reducing to 174 and 57 in female- and male-specific analyses, respectively (**eFigure 5** and **6**).

**Figure 5.**
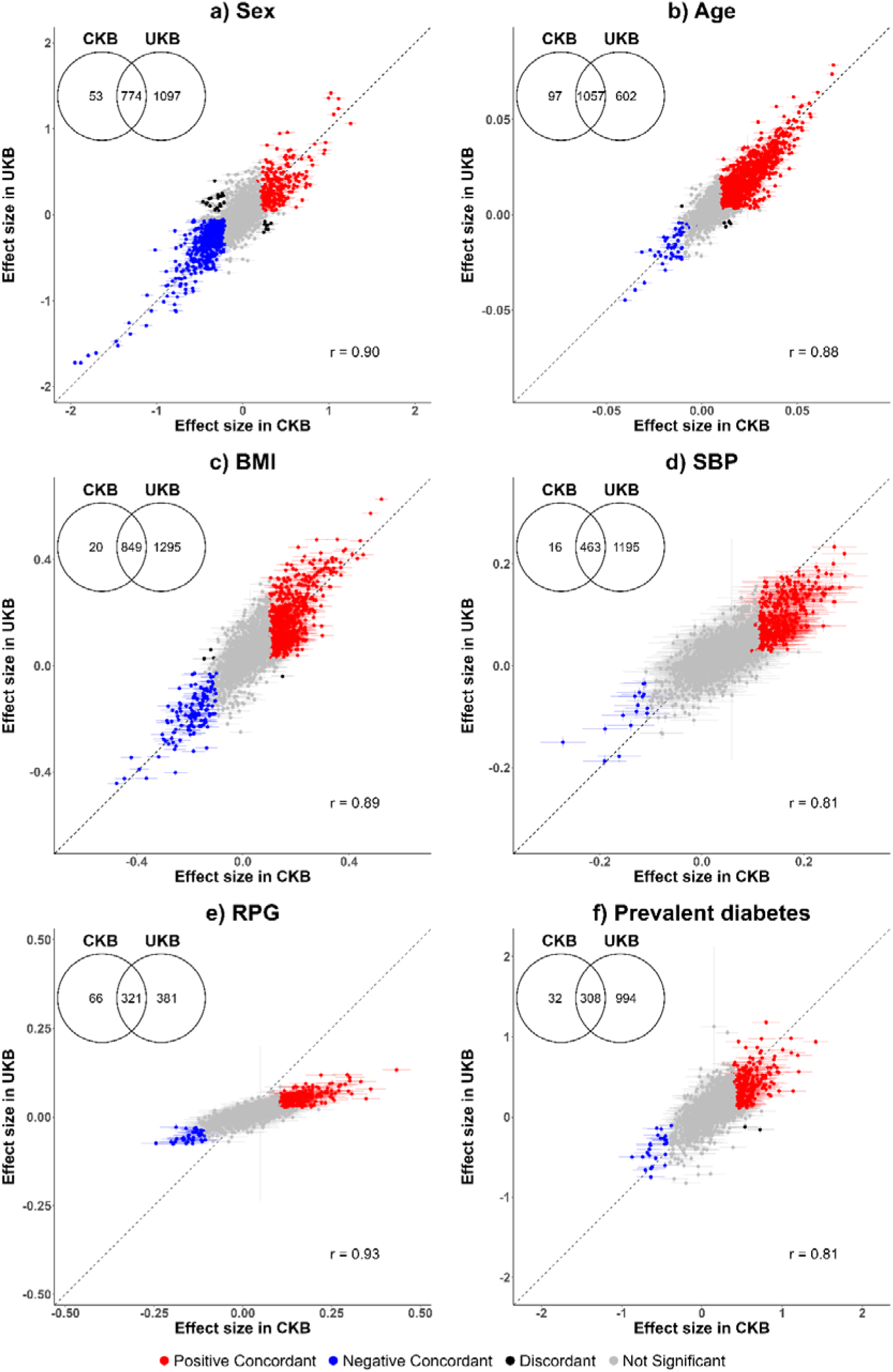
Associations of six selected key baseline characteristics with protein biomarkers in CKB and UKB. In CKB, analyses were adjusted for age, age^2^, sex, study area, fasting time, fasting time^2^, outdoor temperature, outdoor temperature^2^ and plate ID, where appropriate. In UKB, analyses were adjusted for age, age^2^, sex, assessment centre, fasting time, fasting time^2^, season, and plate ID, where appropriate. Abbreviations: BMI: Body mass index; CKB: China Kadoorie Biobank; SBP: systolic blood pressure; UKB: UK Biobank; RPG: Random plasma glucose

Among clinical measurements, BMI was associated with the largest number of proteins (869) followed by SBP (n=479) and RPG (n=387; with 274 overlapping with prevalent diabetes; **Table 1**). For BMI, the strongest associations were observed for LEP, FABPA and IGFBP2 (**eFigure 7a**). In sex-specific analyses, BMI was associated with 576 proteins in females and with 353 proteins in males, with similar strength of associations among the 300 overlapping proteins (r=0.97; **Table 1** and **eFigure 4b**). The leading BMI-related proteins demonstrated similar exposure profiles, with >90% of them also associated with age, sex, prevalent diabetes, and other clinical measurements in both overall and sex-specific analyses (**eFigure 7b, eFigure 8 I.b & II.b**).

Overall prevalent diabetes and HBsAg positivity were associated with 340 and 282 proteins, respectively (**Table 1**), with more associations in women than in men (217 vs. 39 and 198 vs. 45, respectively; **Table 1** and **eFigure 6h)**. Other health-related measures, including self-rated health and prior cancer, were associated with fewer than three proteins. For females, post-menopause was associated with 181 proteins, with the strongest positive association being with FSHB, CGA and DPP4 and the strongest inverse association with PAEP, CHRDL2 and SDK2 (**eFigure 9a**). Additionally, the top menopause-related proteins were predominantly associated with age (**eFigure 9b**).

Healthy lifestyle index was associated with 342 proteins (e.g., IGSF9, VWA1 and LEP) in overall analyses, and with 126 and 99 proteins in male- and female-specific analyses (**Table 1**, **Figure 4 I.a**, **eFigure 10 I.a** & **10 II.a**). Most of these proteins were also associated with the individual components of the index, particularly age, sex and clinical measurements, with alcohol and smoking being particularly notable among males (**Figure 4 I.b**, **eFigure 10 I.b** & **10 II.b**).

Overall the frailty index was associated with 597 proteins (e.g., IGSF9, CDHR2 and IL1RN), with 300 (50%) proteins overlapping with the lifestyle index, albeit in opposite directions in their associations (r=-0.91; **Table 1**, **Figure 4c II.a**). In sex-specific analyses, 102 and 279 proteins were associated with frailty index in females and males, respectively (**eFigure 12 I.a** & **12 II.a**). Many of the frailty index-related proteins were also associated with individual components of the index, including age, sex, clinical measurements, and prior diseases, with similar association patterns in men and women (**Figure 4 II.b**, **eFigure 11 I.b** & **11 II.b**).

In UKB participants (mean age 57 [SD 8.1], 55% female), a total of 1,871 proteins were significantly associated with sex, 1,659 with age, 2,144 with BMI, 1,658 with SBP, 702 with RPG, and 1,302 with prevalent diabetes (**Figure 5**). Over 90% of the significant associations in CKB were replicated in UKB, with the exception of RPG, which had a lower (∼80%) replication rate. However, >95% of RPG associations in CKB were replicated in UKB using HbA1c (**eFigure 12**). Moreover, for the overlapping significant proteins in CKB and UKB, there were high correlations (r>0.80) in effect sizes, with over 95% being directionally consistent. In sex-specific analyses, there were similar patterns and replication rates between the two populations (**eFigure 13**).

## Discussion

In this exposome-wide analysis of Olink proteins in Chinese adults, we identified a large number of associations between various exposures and levels of ∼3,000 proteins. In particular, age, sex and BMI each showed significant associations with plasma levels of >800 proteins. A range of other exposures including, socio-demographic, environmental factors, clinical measurements, health-related traits, and composite lifestyle and health indices, were also associated with levels of modest numbers of proteins. Many proteins were associated with multiple exposures, with CDHR2, CK-BB, and PLAT showing the largest number of associations with the exposures examined. We also observed differences in proteomic-exposure associations between females and males, and replicated >90% of proteomic associations with key exposures in the European populations.

Among all exposures investigated, age yielded the largest number of significant associations with plasma protein levels. The single protein most strongly and positively associated with age was EDA2R, a member of the tumour necrosis factor receptor superfamily.^28^ EDA2R is a known marker for ageing, and its gene expression has been previously reported to be associated with ageing in plasma, muscle, and lung tissues.^29–31^ Moreover, we found that older age was associated with higher levels of ELN, a protein making up elastic fibres in various human organs, including the skin, heart, and blood vessels.^32–35^ Consistent with this finding, we also found a positive association between older age and higher levels of LTBP2, a component of micro-fibrils that interact with ELN.^36^ Finally, many age-related proteins were also significantly associated with other exposures. For example, higher levels of CXCL17, a protein involved in respiratory diseases,^37,38^ were associated with older age, smoking, and amount of exhaled CO in our study.

Our analysis demonstrated marked sex differences in plasma levels of many proteins. These included proteins that are known to be involved in human reproductive processes, such as FSHB which regulates follicular growth in females,^39,40^ as well as ACRV1, EDDM3B, and TEX101, which are involved in spermatogenesis in males.^41–43^ We replicated the well-known finding of elevated LEP levels in females, a protein released by adipocytes and related to sex differences in body fat percentage/distribution.^8,10,11^ Moreover, we found elevated levels of the XG protein in females, an antigen that defines the Xg blood group.^44,45^ Historically, the Xg blood group has received less research attention than other blood groups. However, given its strong associations with sex, age, and BMI in our study, more research is needed to re-evaluate the role of XG in health and disease aetiology. Many of the sex-associated proteins were also independently associated with other exposures, including FSHB, which was also associated with age. Additional analysis revealed that the association between FSHB and age was primarily driven by female sex, since elevated FSHB levels are an indicator of menopause and are commonly observed in post-menopausal women.^46,47^

Our analyses also replicated a number of exposure-protein associations previously reported in Europeans. For example, we identified associations between BMI and 869 proteins, including known associations of higher BMI with higher levels of LEP (regulating energy balance)^48^ and FABP4 (lipid transporter in adipocytes).^49^ Other examples of proteomic associations with clinical measurements include higher SBP and lower REN levels (part of the renin-angiotensin system)^50^ and higher RPG and lower LPL levels (involved in lipid metabolism).^51^ Moreover, several behavioural factors may also affect plasma levels of specific proteins. For example, smoking was associated with higher CXCL17 levels (involved in immune responses and respiratory diseases),^37,38^ and alcohol drinking was associated with higher CHI3L1 levels (involved in liver diseases).^52^ Additionally, factors related to sample collection such as outdoor temperature and fasting time (time since last meal) were also significantly associated with 292 and 79 proteins, respectively in our study. Those included the associations between higher outdoor temperature and higher SPINK6 levels (maintaining skin homeostasis and restricting influenza virus activation)^53,54^ and shorter fasting time and high GIP levels (stimulating insulin secretion).^55^ Such factors related to sample collection could act as potential confounders in associations between exposures and proteins. Indeed, we observed changes in results before and after including them as covariates in the models (**Supplementary eTable 5**), providing justifications for including them in the main models. Given their importance, future studies should also consider collecting such information to improve the robustness and reliability of analyses.

In addition to individual exposures, we also demonstrated novel associations between many plasma proteins and two composite measures that reflect general lifestyle and health, namely the healthy lifestyle index and frailty index. The two indices showed opposing directions of associations with some proteins, including IGSF9, IGFBP2, and SHBG. Both IGSF9 and IGFBP2 have been implicated in multiple types of cancers, and thus have been considered potential diagnostic/prognostic markers and treatment targets.^56–61^ SHBG is a liver-produced secreted protein that binds sex hormones,^62^ which is associated with metabolic and reproductive system disorders.^63–66^ As expected, many proteins associated with the two indices also showed associations with individual exposures, especially BMI, SBP, and RPG. Furthermore, we observed sex differences in proteomic associations for the two indices. For example, the healthy lifestyle index showed stronger inverse associations with GGT1 (a marker of liver function)^67,68^ and CXCL17 (involved in lung function)^37,38^ in males than females. Such difference may be due to the much higher prevalence of alcohol drinking and smoking in males than females (37.2% vs 2.6% and 63.3% vs 2.3%, respectively) in CKB and the general Chinese population.^69–71^

Apart from being the first such study in an East Asian population, the main strengths of the present study include a large number of proteins assayed and a wide range of exposures considered simultaneously in the analyses. Moreover, we also examined potential sex differences in the protein-exposure associations, revealing potential novel findings to inform future research. Nevertheless, the present study also had limitations. First, although our study represented the largest to date amongst proteomics studies in East Asians, it lacked the power for more detailed subgroup analyses beyond sex differences. There were also fewer males than females in our study, which might explain the smaller number of significant associations in males. Second, due to the cross-sectional study design, we could not confirm the direction of the observed associations. Third, although we have adjusted for key covariates (e.g. age, sex, region, outdoor temperature, and fasting time) in our analyses to minimise residual confounding, we could not establish reliably the cause-effect nature of the observed associations, which should be assessed in future studies using genetic approaches (e.g., Mendelian Randomisation). Finally, due to the lack of similar datasets with proteomics data available, we were unable to replicate our findings for all exposures in independent cohorts in Chinese or East Asian populations. However, since over 90% of the associations with sex, age, BMI, SBP and diabetes-related exposures were replicated in UK Biobank, we believe our findings using the same Olink platform should be generalisable to other study populations.

Overall, the present study in Chinese adults demonstrated a large number of proteomic associations across a diverse range of exposures, particularly sex, age, adiposity, and healthy lifestyle and frailty indices. We also identified sex differences in proteomic associations with various exposures, mainly reflecting differences in reproductive processes and lifestyle habits between females and males. Future studies from diverse cohorts are required to replicate our findings and confirm the causal relevance of these associations.

## Abbreviations

ACP5: Tartrate-resistant acid phosphatase type 5
ACRV1: Acrosomal protein SP-10
ALPP: ALPP alkaline phosphatase, placental
BMI: Body mass index
CDHR2: Cadherin-related family member 2
CGA: Glycoprotein hormones alpha chain
CHI3L1: Chitinase-3-like protein 1
CHRDL2: Chordin-like 2
CKB: China Kadoorie Biobank
CK-BB: Brain-type creatine kinase also known as creatine kinase B-type
CRISP2: Cysteine-rich secretory protein 2
CSF3: Granulocyte colony-stimulating factor receptor
CTSV: Cathepsin V
CVD: Cardiovascular disease
CXCL17: C-X-C motif chemokine 17
DPP4: Dipeptidyl peptidase-4
DPT: Dermatopontin
EDA2R: Tumor necrosis factor receptor superfamily member 27
EDDM3B: Epididymal secretory protein E3-beta
EGFR: Epidermal growth factor receptor
ELN: Elastin
Exhaled CO: Exhaled carbon monoxide
FABP4: Fatty acid-binding protein, adipocyte
FGF5: Fibroblast growth factor 5
FSHB: Follitropin subunit beta
GDF15: Growth/differentiation factor 15
GGT1: Glutathione hydrolase 1 proenzyme
GIP: Gastric inhibitory polypeptide
HbA1c: Hemoglobin A1c
HBsAg: Hepatitis B virus surface antigen
HSPB6: Heat shock protein beta-6
IGFBP2: Insulin-like growth factor-binding protein 2
IGFBP3: Insulin-like growth factor-binding protein 3
IGSF9: Protein turtle homolog A
IHD: Ischaemic heart disease
IL17D: Interleukin-17D
IL1RN: Interleukin-1 receptor antagonist
INSL3: Insulin-like 3
ITGB5: Integrin beta-5
ITIH4: Inter-alpha-trypsin inhibitor heavy chain H4
LAMP3: Lysosome-associated membrane glycoprotein 3
LEP: Leptin
LPL: Lipoprotein lipase
LRP1: Low-density lipoprotein receptor-related protein 1
LTBP2: Latent-transforming growth factor beta-binding protein 2
MAMDC4: MAM domain-containing 4
MELTF: Melanotransferrin
NPX: Normalized Protein eXpression
PAEP: Glycodelin
PLAT: Plasminogen activator, tissue type
PON2: Serum paraoxonase/arylesterase 2
pQTLs: Protein quantitative trait locus
QC: Quality control
REN: Renin
RPG: Random plasma glucose
SBP: Systolic blood pressure
SD: Standard deviation
SDK2: Protein sidekick-2
SHBG: Sex hormone-binding globulin
SNED1: Sushi, nidogen and EGF-like domain-containing protein 1
SPINK 6: Serine protease inhibitor Kazal-type 6
TEX101: Testis-expressed protein 101
TIMP4: Metalloproteinase inhibitor 4
VWA1: Von Willebrand factor A domain containing 1
XG: Glycoprotein Xg

## Acknowledgements

The chief acknowledgment is to the participants, China Kadoorie Biobank project staff, staff of the China CDC and its regional offices for access to death and disease registries. The Chinese National Health Insurance scheme provided electronic linkage to all hospital admissions. The China Kadoorie Biobank study is jointly coordinated by the University of Oxford and the Chinese Academy of Medical Sciences.

## Open Access Statement

For the purpose of Open Access, the author has applied a CC-BY public copyright license to any Author Accepted Manuscript version arising from this submission.

## Ethics approval

The China Kadoorie Biobank complies with all the required ethical standards for medical research on human subjects. Ethical approvals were granted and have been maintained by the relevant institutional ethical research committees in the UK and China.

## Consent to participate/publication

All participants provided written informed consent.

## Author Contributions

AI, BW, RC, DAB, ZC conceived and designed the study. AI conducted the statistical analyses and AI and BW wrote the first draft of the manuscript. LL, and ZC as the members of China Kadoorie Biobank Steering Committee, designed and supervised the overall conduct of the study, including obtaining funding for the study. All other authors provided critical revision to the manuscript for important intellectual content. AI, BW, DAB and ZC are the guarantors of this work and take responsibility for the integrity and accuracy of the data analysis. DAB and ZC supervised the work.

## Data Availability

Data from baseline, first and second resurveys, and disease follow-up are available under the China Kadoorie Biobank Open Access Data Policy to bona fide researchers. Details of the China Kadoorie Biobank Data Sharing Policy are available at www.ckbiobank.org.

